# Temporal trends, SARIMA forecasting of Dengue, and the influence of Dengue-related meteorological factors in Bangladesh: a time series analysis

**DOI:** 10.1101/2025.04.09.25325511

**Authors:** Kazi Estieque Alam, Md Jisan Ahmed, Ritu Chalise, Md Abdur Rahman, Tasnia Thanim Mathin, Md Ismile Hossain Bhuiyan, Prajwal Bhandari, Delower Hossain

## Abstract

Dengue is a viral disease spread by mosquitoes and is found primarily in tropical and subtropical areas. Currently, dengue fever (DF) remains a significant public health challenge in Bangladesh, and various meteorological factors influence its incidence. This study aims to analyze the temporal patterns of dengue cases from January 2008 to November 2024 and explore the relationships between meteorological factors and the incidence of dengue fever (DF) in Bangladesh, utilizing time series forecasting models and multivariate Poisson models based on monthly dengue case data. Seasonal decomposition was measured using LOESS seasonal, trend, and residual components. A SARIMA forecast model for dengue cases and Poisson regression assessed meteorological impacts, considering one- and two-month lags. The result indicates that the highest number of dengue cases were found in August 2019 (52,636 cases) and in September 2023 (79,598 cases) with September standing out as the peak month in Bangladesh. Autocorrelation analysis revealed strong positive correlations at 1-month and 2-month lags, indicating the selection of the SARIMA (2,1,2) (1,1,1) [6] model, which effectively captured seasonality with a Mean Absolute Error coefficient of 1649 and a Root Mean Squared Error (RMSE) coefficient of 5203.44. Forecasts from 2024-2027 predict that dengue cases will fluctuate between 10,000 and 20,000 annually. Spearman’s rank correlation indicated positive associations between dengue cases and precipitation (r = 0.37, p<0.05), temperature (r = 0.28, p<0.05), wind speed (r = 0.25, p<0.05), and humidity (r = 0.18, p<0.05). Multivariable Poisson regression revealed that temperature (°C) (IRR = 1.02, 95% CI: 1.02– 1.02, p < 0.001), Humidity (%) (IRR = 1.25, 95% CI: 1.24–1.25, p < 0.001), Wind speed (m/s) (IRR = 1.10, 95% CI: 1.09–1.10, p < 0.001) significantly increased dengue incidence. In conclusion, this study emphasizes the critical role of humidity and temperature in shaping dengue incidence in Bangladesh, highlighting the need to integrate climate data into public health strategies for improved forecasting and control.

## Introduction

Dengue is the most significant mosquito-borne viral disease and is found mainly in tropical and subtropical areas. However, due to globalization and climate change, it has spread to countries where it was not common before (1, 2). This disease is caused by four closely related viruses but antigenically distinct virus serotypes (DEN-1--4) of the genus Flavivirus and is transmitted primarily by the *Aedes (A.) aegypti*, the primary urban vector, and *A. albopictus,* the secondary vector (3). Almost 100 nations in Africa, the Americas, Southeast Asia, the Eastern Mediterranean, and the Western Pacific now have dengue as an endemic disease (4). More than 50% of the global population is at risk of contracting dengue, with an estimated 50 to 200 million cases annually(5). Severe dengue results in approximately 12,500 fatalities each year, primarily in endemic regions where nearly 2.5 billion people are affected (6).

Numerous interconnected elements, including viral serotypes and climatic, hydrological, and societal factors, influence the spread of dengue (7). Rainfall, temperature, and humidity are examples of meteorological parameters that are closely linked to dengue outbreaks. For instance, it has been demonstrated that higher temperatures and rainfall increase the likelihood of dengue infection by increasing the number of mosquito-breeding sites, improving the mosquito breeding cycle, encouraging vector development (shortening the maturation period), increasing vector survival, and promoting blood-feeding patterns. As a result, there are more vectors in the population, which increases the likelihood that a vector and a human would interact and spread the virus (8, 9). The *Ades* mosquito has four stages in its life cycle: the eggs, larvae, and pupae are aquatic, and the adult mosquito is terrestrial. Climate variables such as temperature and rainfall affect each stage of the life cycle (10, 11). Research has shown that *A. albopictus* cannot develop at all at a constant temperature of 10°C and that eggs cannot develop at 15°C. However, when the temperature increased to 30°C, the rate at which each stage developed increased steadily, with the fastest development occurring for the larvae and pupae at approximately 30°C (12). In addition, relatively high temperatures make it possible for mosquito larvae or adults to hibernate (13–16). The endemic range of DF is expected to spread regionally as long as global temperatures rise (17–19). Increased reproduction and activity and a shorter larval incubation period are all made possible by warmer temperatures, which increase the ability to produce progeny. Accordingly, it appears likely that the prevalence and potential for dengue transmission will increase (8, 20). By lengthening the season during which transmission occurs, rising temperatures may also contribute to an increase in dengue transmission (21). Extended dry spells in endemic regions lacking a reliable source of drinking water could promote water storage, which would increase the number of mosquito breeding grounds used by the *A. aegypti* vector (22).

Consequently, Bangladesh has experienced dengue outbreaks in recent years that have never been severe. Furthermore, Bangladesh is close to nations where dengue is endemic, it is constantly in danger of importing novel strains of the disease (23). The deadlier secondary dengue infection may have resulted from the introduction of a new serotype through antibody-dependent enhancement (ADE) (24, 25). Over the past ten years, a growing amount of research has been conducted to anticipate infectious diseases, specifically dengue and malaria. Traditional techniques, which look for a linear association between underlying causes and disease occurrence, such as regression (26) or autoregressive integrated moving average (ARIMA) (27), still, achieve a limited level of accuracy. Moreover, contemporary methods such as support vector machines (SVMs) (28), artificial neural networks (ANNs) (29, 30), and hidden Markov models (HMMs) (31) have been developed.

In Bangladesh, only a few studies have been conducted on the relationship between meteorological factors and the prevalence of dengue, with most of these studies focused on specific regions, particularly Dhaka. The majority of the data used in these studies are relatively old, limiting the scope for understanding recent trends in dengue outbreaks across the country. This regional and temporal limitation highlights the need for more comprehensive research that covers a wider geographic area and includes updated data to better understand how changing climate patterns and weather conditions influence the spread of dengue in different parts of Bangladesh (32–36). As climate change and unusual weather patterns are changing in Bangladesh, dengue has an emerging effect, and few studies have been conducted on the proper relationships between meteorological factors. This study aims to analyze temporal trends in dengue incidence in Bangladesh and forecast future outbreaks until 2027 using the SARIMA model. Additionally, it examines the influence of dengue-related meteorological factors: temperature, rainfall, humidity, and wind speed; through multivariate Poisson regression.

## Materials and methods

### Data collection

For this research study, data on monthly dengue cases were obtained from the Institute of Epidemiology, Disease Control and Research (IEDCR) website (https://iedcr.portal.gov.bd/site/page/45aea1fa-5756-4feb-8d09-f0da895a3baa/-), Dhaka, Bangladesh (37, 38). The IEDCR provides regularly updated public health data, which ensures the accuracy and relevance of the dengue case statistics used in the study. Similarly, meteorological variable measurement data were collected from the Bangladesh Meteorological Department (BMD) (https://bmd.gov.bd/), a reliable platform offering access to a range of environmental and meteorological data of Bangladesh. The data we collected were recorded via the following units: temperature measured in degrees Celsius (°C), rainfall recorded in millimeters (mm), relative humidity expressed as a percentage (%), and wind speed measured in meters per second (m/s).

### Statistical analysis

#### Descriptive

A general descriptive summary and box-and-whisker plots were used to analyze the spread time and determine the dengue case distribution in Bangladesh. A box plot provides a visual representation of the distribution. The box extends from the bottom hinge, which is the 25th percentile, to the higher hinge, which is the 75th percentile. The line across the box represents the median.

#### Decomposition

The seasonal decomposition process based on loess (STL) is used in a variety of study fields, such as the natural sciences, environmental science, and public health, to analyze time series data. SLT separates the long-term, low-frequency variation in the data into three components: trend (the variation in the data over the same period), seasonal (the variation within the same period), and random or remainder (the remaining variation in the data after extracting the trend and seasonal component). The benefits of SLT include its ease of use, reliable outcomes, and potent data visualization. where 𝑌_𝑡_, 𝑇_𝑡_, and R_t_ represents the time series data, trend, seasonal component, and random component, respectively. The following is a description of the equation.

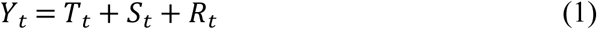

The number of dengue cases in this study is denoted by 𝑌_𝑡_. It is measured on a monthly period. Every year, the number of cases of dengue varies greatly. The numbers during the epidemic year could triple those of the period’s average. As a result, the pattern may be mistranslated. It is imperative to maintain a consistent annual number of dengue cases. As a result, we define a new parameter. 𝑌^∗^, or the corrected dengue data are as follows.

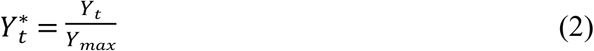

where 𝑌_𝑚𝑎𝑥_ is a dengue case of the peak month of the year. We calculated that the dengue endemic would last six months, from January to December. By lessening the impact of outlier cases, the adjusted value enables us to examine the pattern of dengue incidence.

The variance of *Y_t_*can be described as follows:

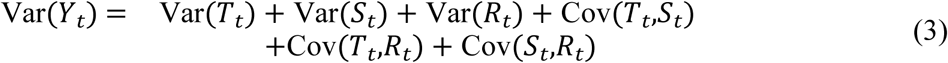

The ratio of the variance of the component and the variance of the dataset was calculated as follows:

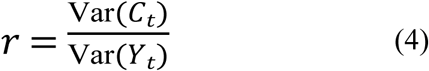

#### SARIMA

An analysis model known as an ARIMA model forecasts potential outcomes via time series data. ARIMA (p, d, q) refers to an ARIMA model that is not seasonal. The number of autoregressive terms, p, is given by the nonnegative integers. The number of times the raw observations are differenced is denoted by d. In the prediction equation, q is the number of lags in the forecast errors. SARIMA (p, d, q)(P, D, Q)m, where m is the number of periods in each season and P, D, Q correspond to the autoregressive, differencing, and moving average terms for the seasonal component, respectively, is an extension of the ARIMA model that incorporates seasonality. (39).

With the assumption that the causes of these events continue to exhibit the same behavior, these models employ nonseasonal differences, autoregressions, and moving average data from prior samples in addition to seasonal differences, autoregressions, and moving averages from prior periods to accurately predict the subsequent steps of a time series.

The mathematical formulation of SARIMA models can be generalized as described in Equation (4).

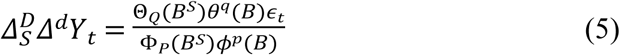

In this equation, *p*, *d,* and *q* represent the nonseasonal order of autoregression (AR), differentiation, and moving average (MA), respectively. P, D, and Q represent the seasonal order of AR, differentiation, and MA, respectively. Moreover, 𝑌_𝑡_ represents the time series data in period *t*. 𝜀_𝑡_ represents the Gaussian white noise process (random walk) in period *t*. *B* represents the backward shift operator 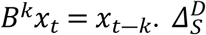 represents the seasonal difference, and 𝛥^𝑑^ represents the nonseasonal difference. *S* represents the seasonal order.

#### Multivariate Poisson regression (MPR)

Since there is usually a lag of months between changes in weather and associated dengue, the goal of this analysis was to determine the best-lagged influence that different meteorological conditions had on dengue occurrence (40). The interval between a single weather observation and a dengue case was used to define the time lag. (41). The monthly meteorological factors and DF cases were correlated for time lags ranging from zero to two months via Spearman’s rank correlation test. The interval between a single weather observation and a dengue case was used to define the time lag. (41).

The following is an example of a basic multivariable Poisson regression model:

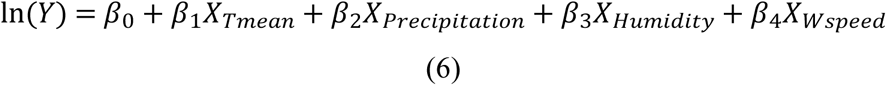

The model adjusted for first-order autocorrelation was as follows:

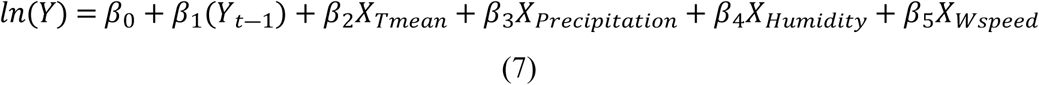

Here, the variables *X_Tmean_*, *X_Precipitation_*, *X_RHumidity_*, and *X_Wspeed_* represent the mean temperature, amount of rainfall, relative humidity, and wind speed, respectively. All the weather variables were tested for multicollinearity with variance inflation factors (VIFs).

### Statistical tools and packages

In this study, the R programming language was employed to handle various statistical tasks. Descriptive statistics and regression model summaries were generated via the ‘*gtsummary’* package, which provides easy-to-interpret, publication-ready tables of results. (42). The ‘*MASS’* package was used to conduct Poisson regression, fitting models for count data (43). Time series analysis and forecasting were performed via the *forecast* and ‘*dlnm’* packages, enabling decomposition and distributed lag models to understand temporal patterns (44). For data visualization, the ‘*ggplot2’* package was used to create sophisticated plots (45), while the ‘*corrplot’* package helps in visualizing the correlation matrix effectively, uncovering hidden relationships among variables (46).

## Results

### Dengue cases and meteorological factors

There are large variations in the number of dengue cases in Bangladesh, with an average of 3,093 and a standard deviation (SD) of 10,960 from January 2008 to November 2024. Meteorological parameters showed a maximum temperature of 30.47°C (SD±3.05°C) and a low of 21.6°C (SD±5.2°C), with a wider range of 1.6-27.5°C. The mean temperature showed moderate variability, averaging 25.7°C (SD±4.1°C). Precipitation and humidity also vary considerably, while wind speed remains relatively stable (**Table 1**).

**Table 1.**
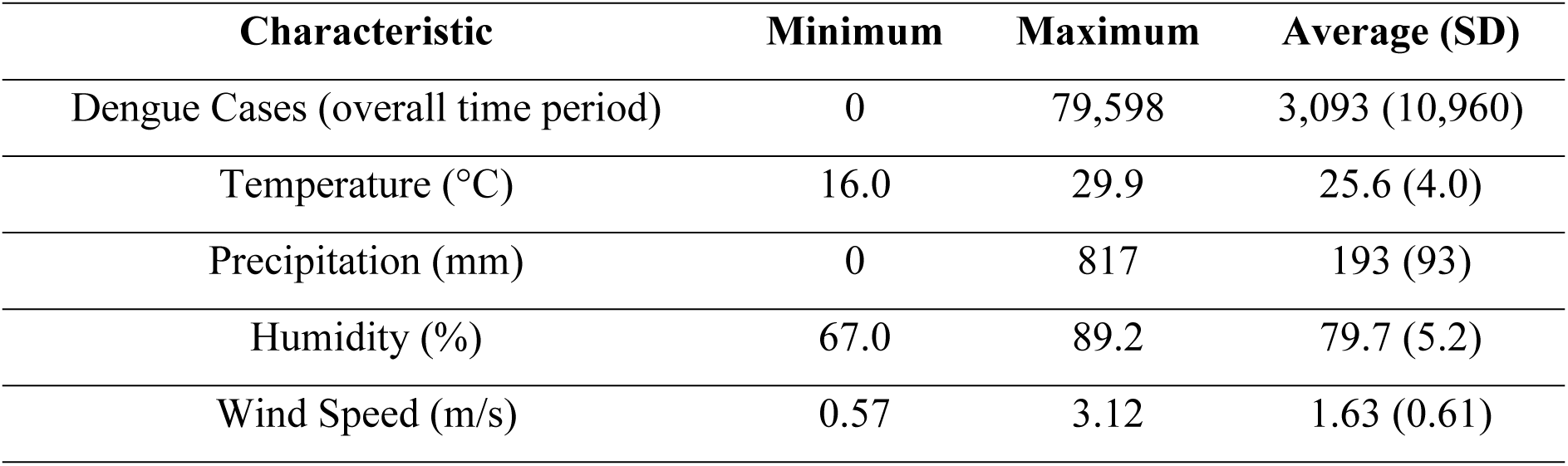
Descriptive statistics of dengue cases and meteorological factors from Jan 2018-Aug 2024 in Bangladesh.

| Characteristic | Minimum | Maximum | Average (SD) |
| --- | --- | --- | --- |
| Dengue Cases (overall time period) | 0 | 79,598 | 3,093 (10,960) |
| Temperature (°C) | 16.0 | 29.9 | 25.6 (4.0) |
| Precipitation (mm) | 0 | 817 | 193 (93) |
| Humidity (%) | 67.0 | 89.2 | 79.7 (5.2) |
| Wind Speed (m/s) | 0.57 | 3.12 | 1.63 (0.61) |

### Temporal distribution of dengue cases

The highest peak number of dengue cases between January 2008 and November 2024 was found in August 2019 (52,636 cases) and in September 2023 (79,598 cases) along with 71,976 cases in October same year, both years experiencing significant peaks surpassing 40,000 cases (**Figure 1**). The boxplot shows that the maximum number of dengue cases is found from July to November, with September standing out as the peak month for most dengue cases in Bangladesh, and these months also contain outliers of the cases (**Figure 2**).

**Figure 1.**
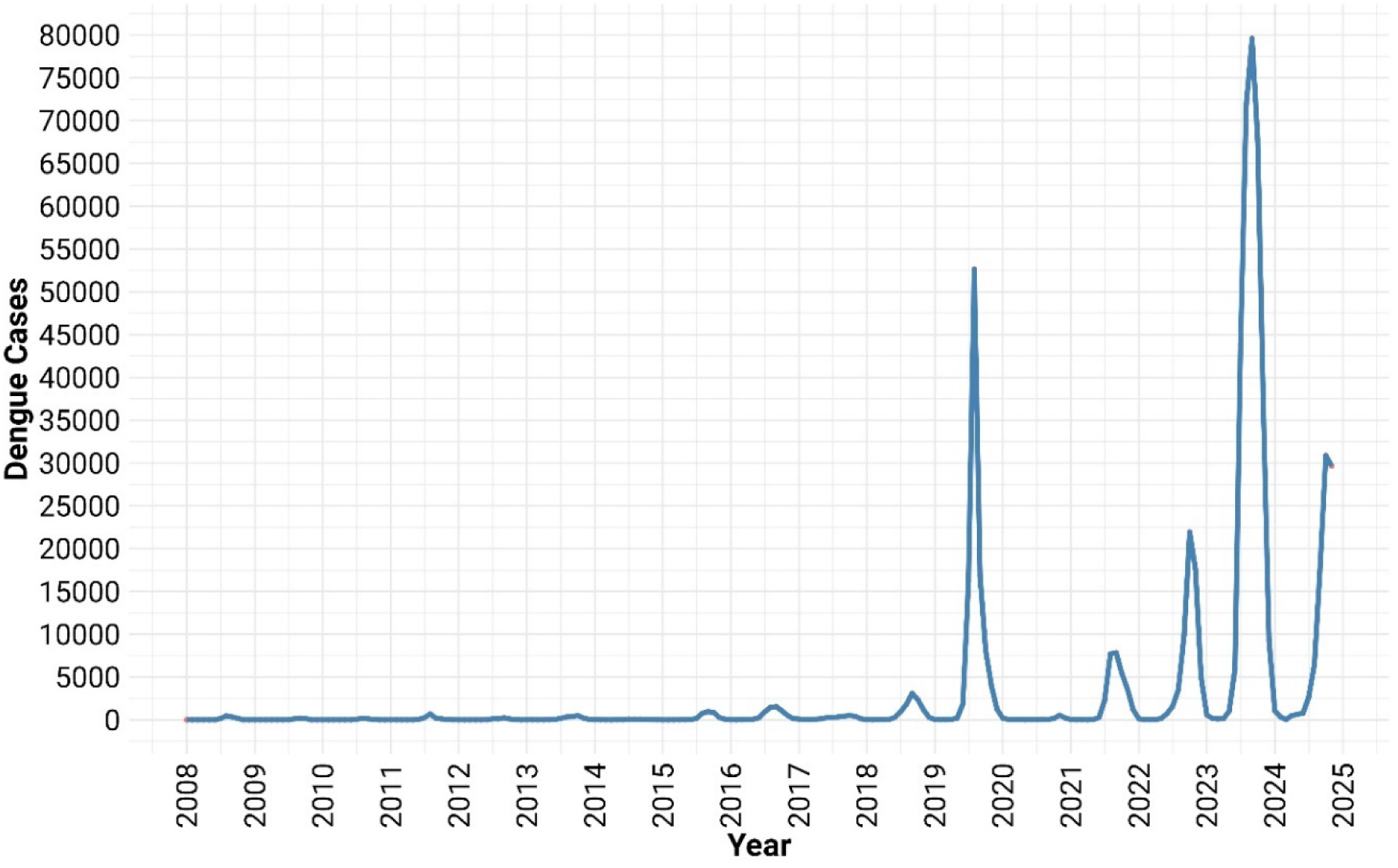
Number of dengue cases from January 2018 to August 2024.

**Figure 2.**
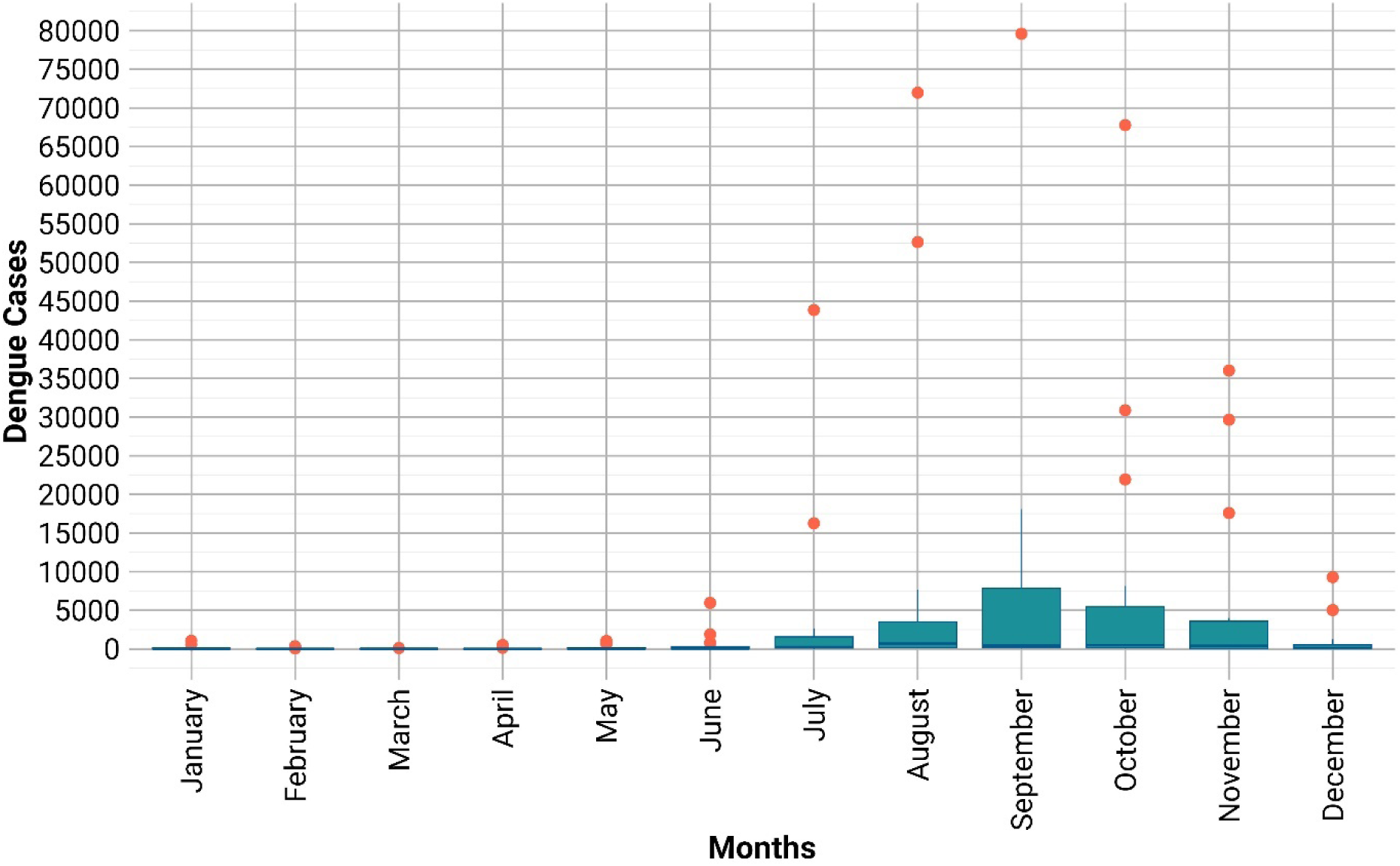
Monthly box plot distribution of the dengue cases Decomposition.

We created the adjusted data from the raw data by using Eqs. (1 and 2). **Figure 3** shows the STL plot of the raw data. There are prominent high peaks in the data, reflecting significant events, whereas the seasonal component captures strong annual cycles with regular peaks, likely indicating periodic factors. The trend component shows a gradual increase followed by a decline, suggesting greater underlying influences on the data. The remainder of the component displays random fluctuations and spikes, which could represent unexpected events or noise after removing the seasonal and trend effects.

**Figure 3.**
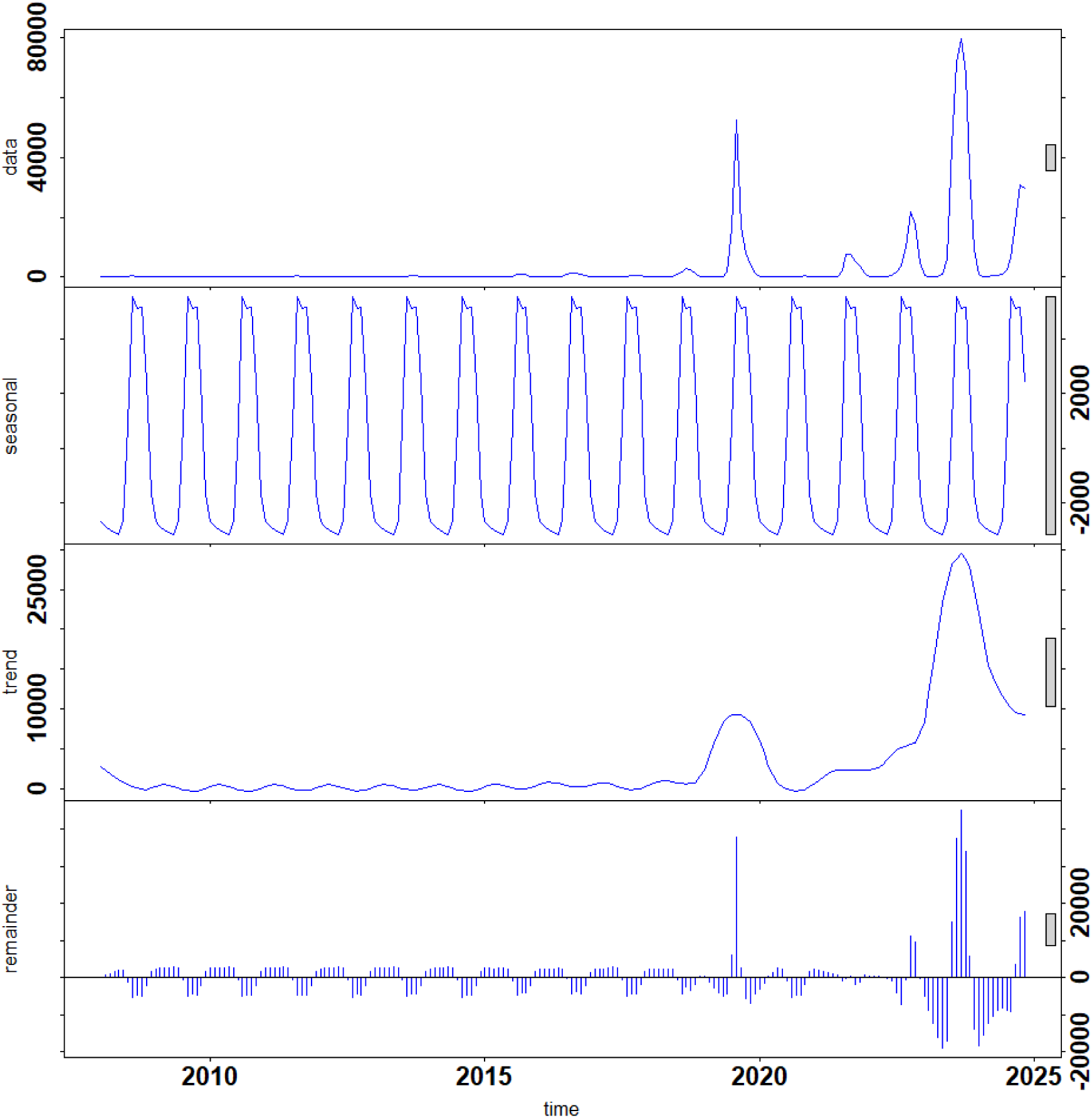
Decomposition plot of the time series of dengue cases in Bangladesh from January 2008 to November 2024.

### Autocorrelation and SARIMA

The autocorrelation (ACF) and partial autocorrelation (PACF) analyses for dengue cases from January 2008 to November 2024 revealed a strong seasonal pattern (**Figure 4**). The ACF plot shows significant positive autocorrelation at lag 1 (∼1 month) and lag 2 (∼2 months), with values around 0.8 and 0.5, respectively, indicating that dengue cases are highly correlated with cases from the preceding months. Additionally, moderate autocorrelation is observed around lags 10–12, suggesting possible annual seasonality. The PACF plot exhibits a large spike at lag 1 (∼1 month) with a value above 0.5, followed by no significant spikes beyond lag 2, implying that the short-term correlation is primarily driven by the most recent monthly cases. The blue dashed lines in both plots indicate significance thresholds, reinforcing the presence of strong autocorrelations within the first few months.

**Figure 4.**
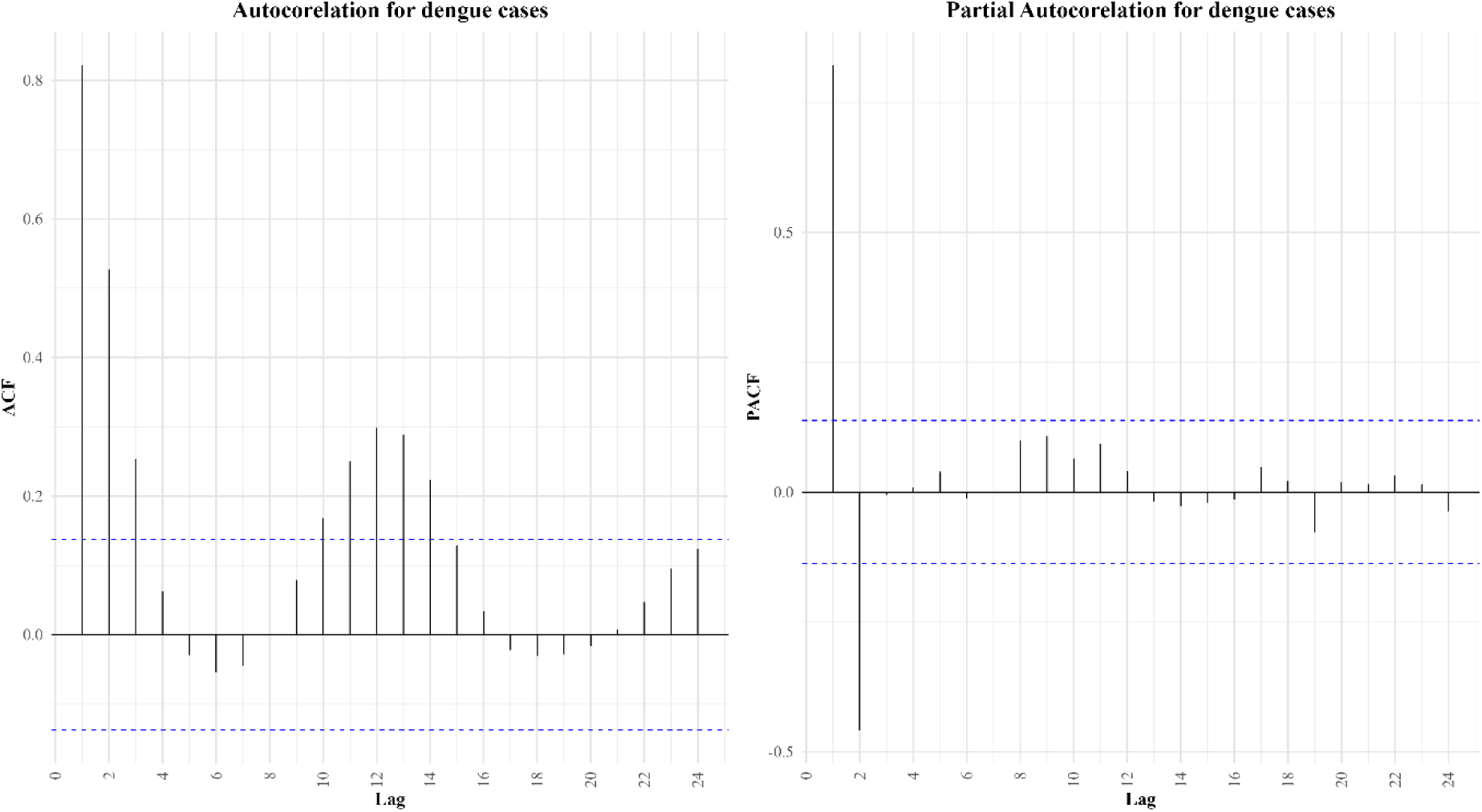
ACF and PACF plots of dengue cases.

SARIMA (Seasonal Autoregressive Integrated Moving Average) model identification involves determining the appropriate orders of autoregression (AR), differencing (I), and moving average (MA) terms for both seasonal and non-seasonal components of the time series. Based on the ACF and PACF plots for dengue cases from January 2008 to November 2024, a first-order differencing (𝑑=1) is needed due to the slow decay of the ACF, indicating non-stationarity. The strong spike at lag 1 in the PACF suggests an AR (1) process (𝑝=1 or 2), while the ACF pattern indicates a moving average component (𝑞=1 or 2). The first-order seasonal differencing (𝐷=1). The seasonal ACF and PACF patterns indicate the need for a seasonal autoregressive term (𝑃=1) and possibly a seasonal moving average term (𝑄=0 or 1). The 6-month lag was constructed and determined based on autocorrelation (**Figure 4**) and the monthly distribution pattern of dengue cases (**Figure 2**) and based on the cumulative effect of environmental conditions on case occurrence. From the ACF and PACF result 14 SARIMA models can be constructed and the best model was selected by checking the best model selection criteria like mean absolute error (MAE), Root Mean Squared Error (RMSE), and L Jung (p-value) (p > 0.05), and the model selection criteria revealed the following results: Root Mean Squared Error (RMSE): 5203.34, and Mean Absolute Error (MAE) = 1649.56 and determined the best model which is SARIMA (2,1,2) (1,1,1) [6] (**Table 2**).

**Table 2.** SARIMA model characteristics for dengue cases.

| SARIMA MODELS | ME | MAE | RMSE | L Jung (*p*-value) |
| --- | --- | --- | --- | --- |
| SARIMA (1,1,1) (1,1,1) [6] | 81.57 | 1740.57 | 5770.859 | 0.030 |
| SARIMA (1,1,2) (1,1,1) [6] | 79.80 | 1732.75 | 5762.692 | 0.044 |
| SARIMA (2,1,1) (1,1,1) [6] | 348.48 | 1622.45 | 5228.877 | 0.952 |
| SARIMA (2,1,2) (1,1,1) [6] | 349.71 | 1649.56 | 5203.34 | 0.987 |
| SARIMA (2,1,2) (2,1,2) [6] | 345.67 | 1660.55 | 5203.984 | 0.971 |

The prediction from 2024-2027 in the SARIMA (2,1,2) (1,1,1) [6] model shows periodic oscillations in the number of dengue cases every six months (**Figure 5**), following a clear seasonal pattern. The predicted cases range between 10,000 and 20,000, with noticeable peaks occurring regularly, suggesting recurring outbreaks. The 95% confidence interval widens as the forecast progresses, indicating increasing uncertainty. Despite this uncertainty, the model predicts consistent seasonal peaks, reflecting the expected cyclical nature of dengue during the forecast period. The negative lower bound in the 95% confidence interval (CI) is a statistical artifact rather than a realistic prediction. ARIMA-based models assume normally distributed errors, leading to symmetrical confidence intervals around the forecasted values. As a result, the lower bound may extend below zero, even though negative dengue cases are not possible. Additionally, transformations like Box-Cox or differencing can further influence the CI range.

**Figure 5.**
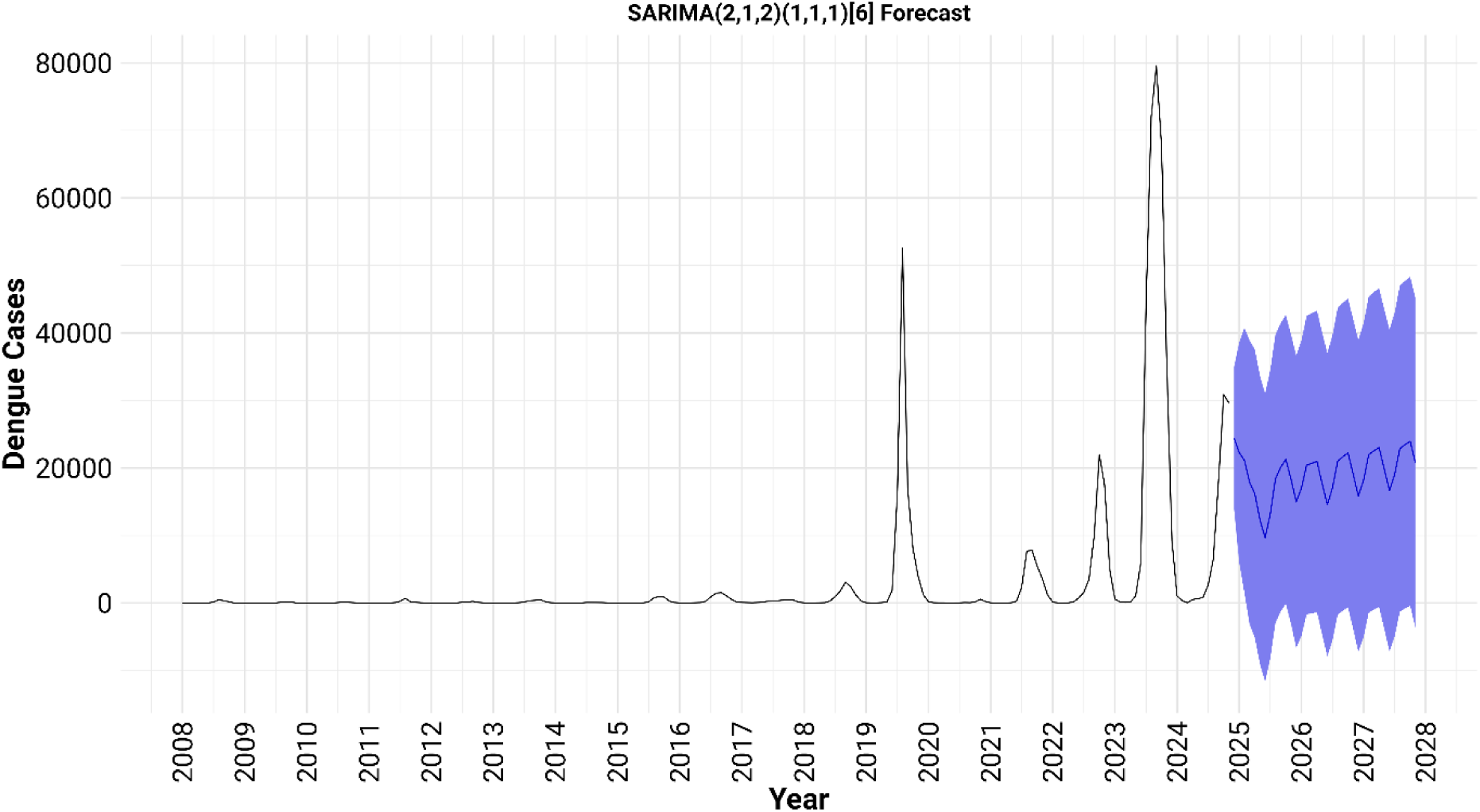
Seasonal SARIMA (2,1,2) (1,1,1) [6] forecast of dengue cases (2024–2027) with 95% confidence intervals.

### Correlation analysis of meteorological factors with dengue cases

Spearman’s rank correlation analysis (**Figure 6**) revealed that dengue cases had the strongest positive correlation with Precipitation (r = 0.37, p<0.05), indicating that as rainfall increased, so did dengue cases. Temperature (r = 0.28, p<0.05) and wind speed (r = 0.25, p<0.05) also exhibited positive correlations with dengue cases which also indicated the increase in dengue. The relationship between dengue cases and humidity (r = 0.18, p<0.05) is weaker but has a significant relation.

**Figure 6.**
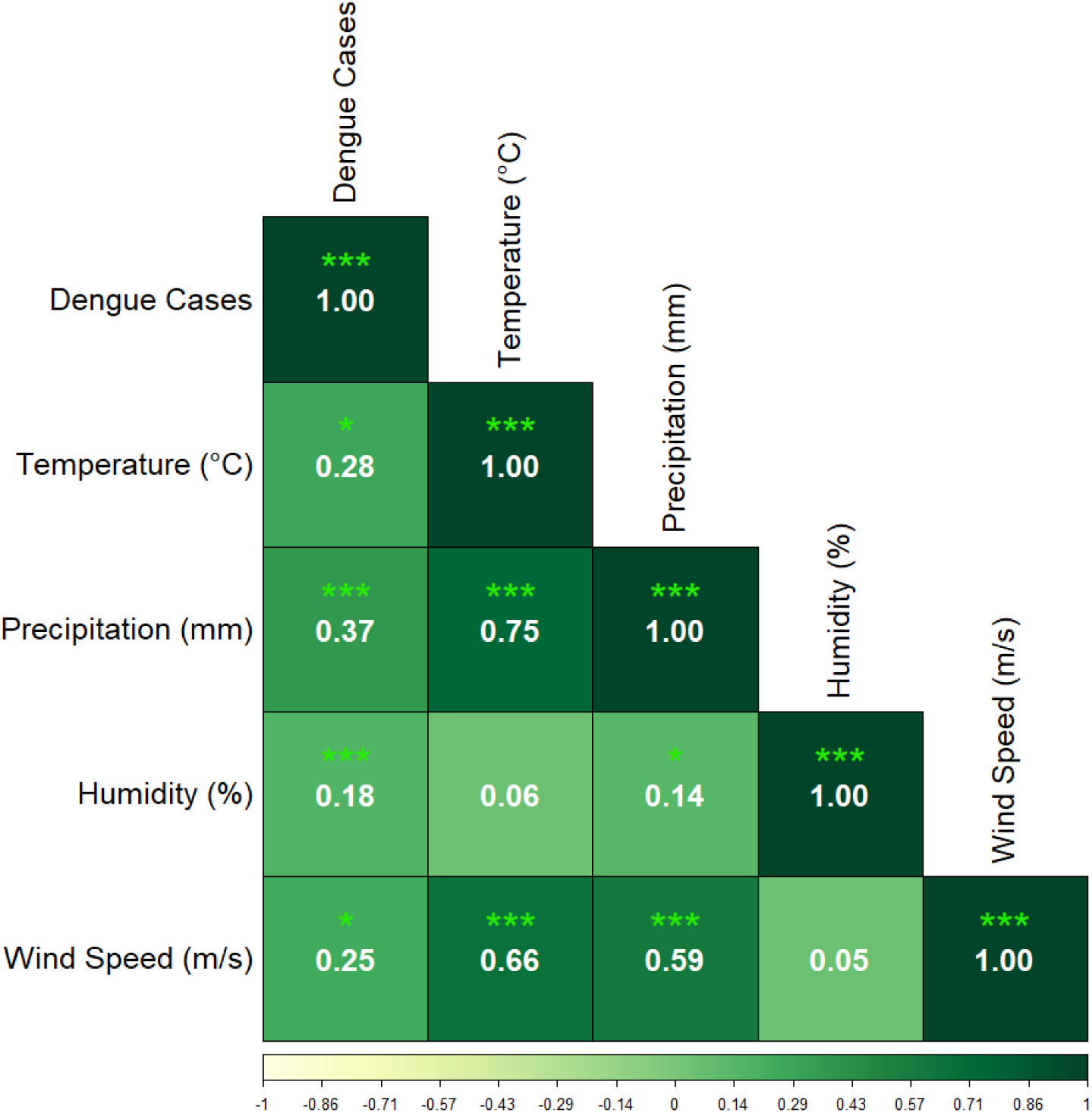
Spearman’s rank correlation plot of the associations of dengue cases with meteorological factors.

The temperature, precipitation, and humidity exhibited significant positive correlations with dengue cases at a one-month lag (**Figure 7**). Specifically, temperature (°C) lag1 showed a moderate positive correlation with dengue cases (r = 0.49, p < 0.001). Precipitation (mm) lag1 displayed a stronger correlation (r = 0.56, p < 0.001), while humidity (%) lag1 had a weaker positive association (r = 0.17, p < 0.001). Wind speed (m/s) lag1 also showed a moderate positive correlation (r = 0.47, p < 0.001). With a two-month lag, the associations strengthened. Temperature (°C) lag2 demonstrated a higher correlation with dengue cases (r = 0.60, p < 0.001), and precipitation (mm) lag2 maintained a strong positive relationship (r = 0.60, p < 0.001). Humidity (%) lag2 remained weakly associated (r = 0.13, p < 0.05), while wind speed (m/s) lag2 showed a stronger correlation (r = 0.65, p < 0.001). These findings indicate that temperature and precipitation play a crucial role in influencing dengue outbreaks with a time-lag effect. (**Figure 7**).

**Figure 7.**
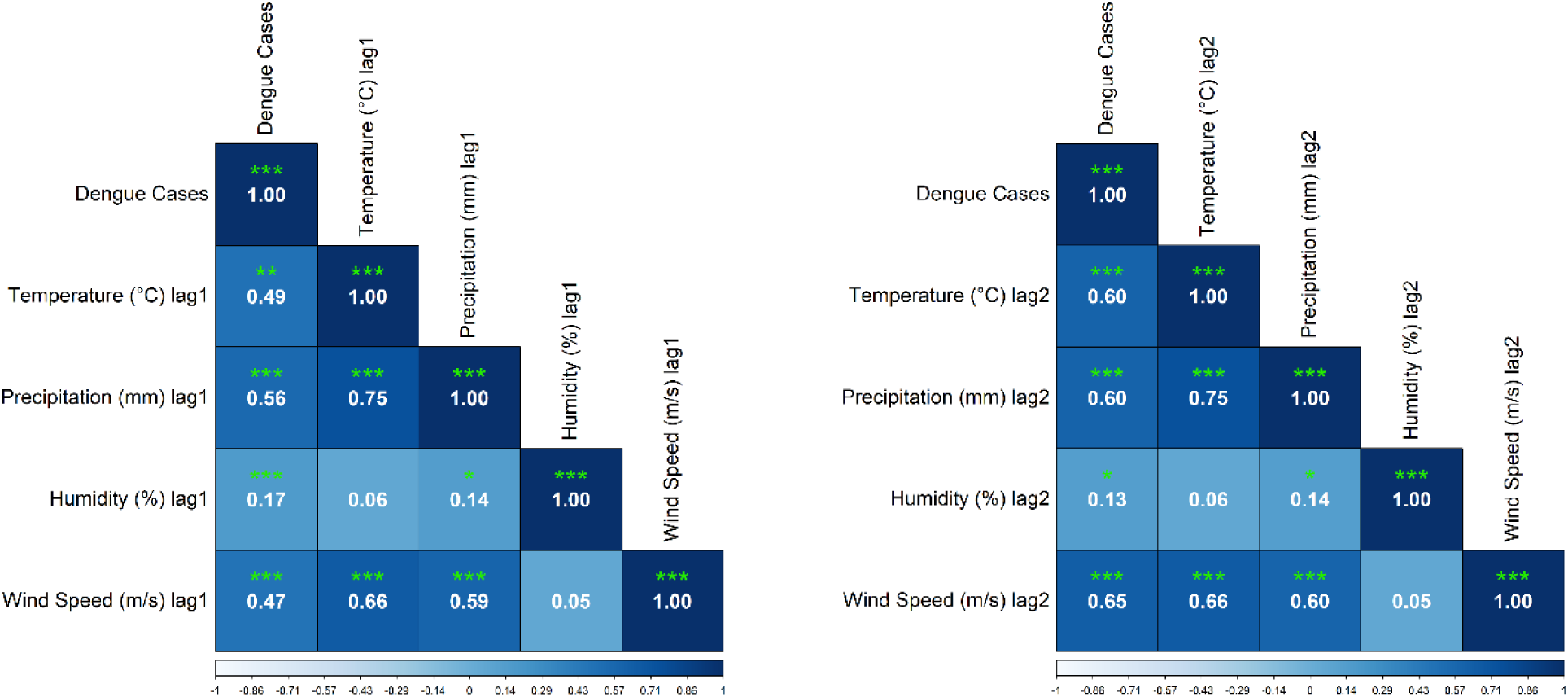
Spearman’s rank correlations between meteorological variables and dengue cases in Bangladesh with one month and two months of lag.

### Multivariable Poisson regression

The Poisson regression results indicate that various weather factors have significant effects on the cases of dengue (**Table 3**). An increase in temperature (°C) (IRR = 1.02, 95% CI: 1.02– 1.02, p < 0.001) is associated with an increase of 2% cases of dengue, likely due to the acceleration of mosquito development and increased virus replication under warmer conditions. Precipitation (mm) (IRR = 1.00, 95% CI: 1.00–1.00, p < 0.001) shows a neutral effect, suggesting that while rainfall provides breeding sites, excessive precipitation may also wash away mosquito larvae. Humidity (%) (IRR = 1.25, 95% CI: 1.24–1.25, p < 0.001) increases dengue cases by 25%, likely because higher humidity enhances mosquito survival, feeding behavior, and overall vector activity. Wind speed (m/s) (IRR = 1.10, 95% CI: 1.09– 1.10, p < 0.001) is associated with an increase of 10% dengue cases, indicating that moderate wind speeds may facilitate mosquito dispersal, potentially increasing the reach of infected vectors.

**Table 3.** Time series Poisson regression of the monthly dengue cases (2008-2024) on the weather factors.

| Variables | IRR<sup>I</sup> | 95% CI<sup>II</sup> | p-value |
| --- | --- | --- | --- |
| *Temperature (°C)* | 1.02 | 1.02, 1.02 | <0.001 |
| *Precipitation (mm)* | 1.00 | 1.00, 1.00 | <0.001 |
| *Humidity (%)* | 1.25 | 1.24, 1.25 | <0.001 |
| *Wind Speed (m/s)* | 1.10 | 1.09, 1.10 | <0.001 |

<sup>I</sup>IRR = Incidence Rate Ratio, <sup>II</sup>CI = Confidence Interval

### Multivariable Poisson regression with lagged

Table 4 presents the Poisson regression results with one-month and two-month lags to assess the influence of weather variables on monthly cases in Bangladesh. In the one-month lag model, temperature showed a negative and statistically significant effect on dengue cases (IRR= 0.319, 95% CI: 0.316–0.322, p<0.001), indicating that higher temperatures in the previous month were associated with lower cases. Precipitation also had a significant negative effect (IRR= 0.002, 95% CI: 0.002–0.002, p<0.001), suggesting that increased rainfall may reduce dengue transmission, possibly by flushing mosquito breeding sites. Similarly, humidity (IRR= 0.193, 95% CI: 0.192–0.193, p<0.001) and wind speed (IRR= 0.601, 95% CI: 0.596– 0.606, p<0.001) negatively impacted cases. In the two-month lag model, temperature was still negatively associated with dengue cases but with a weaker effect (IRR= 0.935, 95% CI: 0.930– 0.940, p<0.001). Precipitation (IRR= 0.001, 95% CI: 0.001–0.001, p<0.001) and humidity (IRR= 0.106, 95% CI: 0.105–0.106, p<0.001) continued to show significant negative effects. Wind speed also exhibited a negative association with cases (IRR= 0.933, 95% CI: 0.928– 0.939, p<0.001), indicating that stronger winds may disrupt mosquito activity and reduce transmission risk.

**Table 4.** Time series Poisson regression of the monthly dengue cases (2008-2024) with one- and two-month lags on the weather factors.

| Variables | Poisson regression with one month Lag |  |  | Poisson regression with two-month Lag |  |  |
| --- | --- | --- | --- | --- | --- | --- |
|  | IRR<sup>I</sup> | 95% CI<sup>II</sup> | p-value | IRR<sup>I</sup> | 95% CI<sup>II</sup> | p-value |
| *Temperature (°C)* | 0.319 | 0.316, 0.322 | **0.000** | 0.935 | 0.930, 0.940 | **0.000** |
| *Precipitation (mm)* | 0.002 | 0.002, 0.002 | **0.000** | 0.001 | 0.001, 0.001 | **0.000** |
| *Humidity (%)* | 0.193 | 0.192, 0.193 | **0.000** | 0.106 | 0.105, 0.106 | **0.000** |
| *Wind Speed (m/s)* | 0.601 | 0.596, 0.606 | **0.000** | 0.933 | 0.928, 0.939 | **0.000** |

<sup>I</sup>IRR = Incidence Rate Ratio, <sup>II</sup>CI = Confidence Interval

## Discussion

Dengue development, transmission, and incidence are closely tied to climate. As a vector-borne disease, its spread heavily relies on the presence and abundance of its vector. Numerous studies have shown that ecological and climatic factors significantly affect the seasonal occurrence of the dengue virus (47–52). In tropical and subtropical regions, climate change poses a severe threat to global health by enabling the spread of DF. Bangladesh’s total number of dengue cases has risen steadily since 2010, surpassing 80,000 confirmed cases in 2023.

The dengue outbreak in 2023 was unusually severe, with incidence rates more than ten times higher than the average reported during the study period. This spike could lead to significant errors in predictive models, as it deviates considerably from the typical trend. The link between DF and climatic patterns remains poorly understood because of the complexity of the vector and host life cycles. The life cycle of the *A. aegypti* mosquito, the primary vector for dengue, is highly sensitive to climate conditions, which adds to the challenge of fully understanding this relationship (48, 53). However, previous research conducted in South Asia and Southeast Asia has examined the role of climate change in the spread of DF (47, 48, 54–56). These studies identified temperature, precipitation, wind speed, and humidity as key climatic factors that may contribute to dengue outbreaks. They also demonstrated the consistent impact of certain climate conditions over time. In this research, we assess the effects of climate variables on dengue incidence, providing valuable insights for mitigating severe future outbreaks and enhancing preparedness efforts in Bangladesh.

The fluctuating trends of DF demonstrate that meteorological factors substantially affect dengue occurrence in southern Thailand (57). Our research indicates that dengue transmission in Bangladesh transpired from January to December throughout the years 2018 to 2024. The longest period of dengue transmission in Bangladesh occurred from July to November during the study period. These findings correspond with other studies conducted in southern Thailand (57), which indicated that the most extended dengue transmission season in the Gulf of Thailand occurred from June to September. This aligns with previous findings in Singapore (58), which indicated a rise in dengue incidence from June to October, perhaps because of the geographic and climatic similarities between Thailand and Singapore. Dengue incidence in Delhi, India, peaks in April and from June to October, which is similar to Bangladesh, likely because of the varying monsoon periods between Thailand and India (26). Moreover, India’s peak temperatures may reach 45°C during the summer months of April, May, and June, hence influencing dengue transmission dynamics (26).

In our study, the autocorrelation plot revealed significant positive autocorrelation at the 1-month and 2-month intervals for dengue cases. The virus typically has an incubation period of seven to fourteen days, during which it progresses from an infectious state to an active viral disease. The stability of the dengue virus often remains in endemic equilibrium, influenced by these delay times. Consequently, vector-borne illnesses may persist in carriers for up to one to two weeks (59–61). The autocorrelation function (ACF) plot was truncated at lag 4, which was parameterized to a moving average (MA) model with no seasonal lag observed in Nakhon Si Thammarat (62). In contrast, Islam *et al*. (36) reported that after 12 weeks, the autocorrelation of DF cases gradually decreased to negligible levels. Peaks in the partial autocorrelation were noted at weeks 1 and 4. Chumpu *et al*. (63) also reported that 1-week lag cases had the highest correlation among 65 provinces in Thailand.

In our study, the best-fitting ARIMA model was identified as SARIMA (2,1,2) (1,1,1) [6]. Similarly, Gui *et al*. (64) determined that ARIMA (1,1,0) with first-order differencing and one autoregressive term was the optimal model for capturing the trend and autoregressive properties of the dengue time series. In Bangkok, (39) reported that the best-fitting model was SARIMA (1,0,2)(1,1,2) [12], which has a strong seasonal component (1,1,2) combined with a nonseasonal component (1,0,2), indicating a mixed model. For overall dengue incidence in northeastern Thailand, Silawan *et al*. (65) fitted a seasonal ARIMA (2,1,0) (0,1,1) [12] model to data spanning 1996--2003. Additionally, Earnest *et al.* (66) reported that ARIMA (3,1,0) provided the lowest mean absolute percentage error (MAPE) of 19.86, indicating its effectiveness in their study.

Temperature has been identified as a key predictor for the increase in dengue cases. In this study, a significant correlation was found between dengue incidence from 2008--2024 and factors such temperature, precipitation, windspeed and humidity. These findings align with those of a previous study by Pinto *et al*. (55), which showed that minimum and maximum temperatures are strong indicators for predicting dengue case increases on the basis of data from Singapore between 2000 and 2007.

Temperature is the most crucial climatic factor influencing the growth and spread of mosquito vectors, making it a potential predictor of dengue outbreaks (57). It significantly impacts the entire life cycle and behavior of mosquitoes, including population density, biting rates, gonotrophic cycle durations, and vector size (67, 68). These findings align with earlier research (47, 69), confirming that extreme temperatures negatively affect dengue vector development. As a result, temperature influences vector efficiency and the potential for an epidemic (70). Given the clear link between temperature and dengue incidence, projected temperature changes could intensify dengue transmission in Bangladesh. A possible explanation is that the optimal breeding temperature range for *A. aegypti* mosquitoes is 20-35°C (49). In our study, the minimum and mean temperatures ranged between 21°C and 30°C, providing ideal conditions for mosquito breeding and facilitating the transmission of dengue viruses.

Another key factor is humidity, which increases the number of dengue patients in Bangladesh. The correlation between dengue incidence and humidity was strongly positive. Similar results were also reported in previous studies (65, 71). This may be attributable to the influence of relative humidity on rainfall, as it indicates the degree of moisture saturation in the atmosphere. A reduction in temperature while the air is saturated can induce condensation, ultimately producing rain (50).

This study revealed that precipitation was associated with dengue cases. In our study, the lag 1 and lag 2 correlations were strongly correlated and were crucial influencing factors in dengue endemics. This outcome is supported by previous studies (72, 73). In Bangladesh, the rainy season lasts from April to September (74). Rainfall is known to have both positive and negative effects on mosquito growth. On the one hand, light rainfall provides standing water, which is essential for mosquito breeding. On the other hand, excessive or excessive rainfall can disrupt mosquito development by destroying mosquito breeding sites or washing away larvae, limiting their growth potential (75). In this research, we found that there was a significant increase from July to November, just before the dry season started. Moreover, the peak season for rain is April to June, when the rate is quite low because of extreme rainfall compared with that in later months (48). This study also revealed that wind speed was weakly positively correlated with dengue incidence in Bangladesh, which is similar to the findings of a previous study (48, 76).

In our study, temperature, precipitation, humidity, and wind speed had significant effects on dengue cases between 2008 and 2024. Meteorological factors mostly affect the increase in dengue cases worldwide. The relationships between weather factors and dengue cases may vary across different periods, especially with increasing global warming and urbanization (64). (77) reported that minimum temperature, humidity, and rainfall were key factors influencing dengue cases, whereas maximum temperature and maximum humidity were not significant in the Mekong Delta area in Vietnam.

In the northern region of Thailand, the 1-week lag is the most significant predictor of dengue incidence, except in Phichit and Sukhothai, where 3-week lag is more important. Key weather factors include negative current‒week pressure, 3-week lag wind power, vaporization, and wind direction. The average model accuracy for the region is 0.72, with more dengue cases linked to low pressure and high humidity, as seen in provinces such as Lampang and Tak. The 1-week lag cases are the strongest predictors, except for Si Sa Ket, where the 2-week lag cases are more relevant. The most significant weather variable is 3-week lag vapor, followed by pressure and wind direction. The average accuracy here is 0.67, with low pressure linked to high dengue incidence in provinces such as Nakhon Ratchasima. (63).

One of the primary causes of Singapore’s high degree of dengue expansion over the previous 40 years has been urbanization (78). According to Lee *et al*.’s calculations, Ho Chi Minh City’s dengue transmission model does not significantly depend on any of the climatic variables (79). According to Johansson *et al*. the accuracy of seasonal autoregressive dengue models for Mexico improved slightly when climate data were used (80). In certain districts of Thailand, there is a substantial correlation between the number of dengue cases and the number of female mosquitoes as well as among seasons (81).

Research has shown that temperature, relative humidity, and rainfall all affect the spread of DF, either directly or indirectly (26, 82). Temperature affects the extrinsic incubation period, reproduction rates, and vector population growth (83, 84). Research has indicated that the confluence of temperature and humidity has a major effect on the quantity of blood consumed and increases the vector survival rate (85). Numerous studies have demonstrated that the connection between climatic conditions and dengue transmission varies over time and across the Asia-Pacific region (86, 87). The significance of the lag time of climate factors has also been emphasized by numerous studies (88, 89). For example, there was a substantial positive correlation with daily rainfall at a lag of 10 weeks, the minimum temperature at a lag of 1-3 months, the maximum temperature at a lag of 1-4 months, and the relative humidity at a lag of 1-3 months in Taiwan (90).

One limitation of this study is the exclusion of important socioeconomic and urbanization-related factors that could significantly influence dengue transmission. Variables such as population density, income levels, housing conditions, access to healthcare, and sanitation infrastructure play a crucial role in shaping disease dynamics. Additionally, factors like land use patterns, urban expansion, and climate variations interact with mosquito breeding habitats, further affecting dengue incidence. Incorporating these parameters in future studies could enhance the accuracy and comprehensiveness of dengue risk assessments and predictive models.

According to our research, the incidence rates of minimum temperature, relative humidity, and rainfall in Bangladesh were significantly related to one- and two-month lags, respectively. This is also in line with a study conducted in Taiwan (90). Understanding how climate factors affect *Aedes* mosquito development, maturation, and survival (approximately 7–9 days from egg to adult) as well as how long DENV takes to incubate intrinsically in humans (4–6 days) (91) and extrinsically in the vector (10 days) (92, 93) is important. The duration and humidity of dormancy in the environment can impact larval survival, developmental rates, and the production of relatively small adults. Eggs are resistant to desiccation and can endure months of dormancy (94).

This study has significant ramifications, although it also has considerable shortcomings. The dataset, intended to evaluate the influence of climate variables on dengue incidence in Bangladesh, contains several missing or incomplete records, reflecting inadequate data collection and reporting practices. This may arise from insufficient monitoring mechanisms or a deficiency in awareness among reporting authorities. Consequently, deficiencies in population data may result in biases in estimates due to underreporting or overreporting, given that the dataset encompasses both confirmed and suspected dengue cases.

## Conclusion

This study reveals significant insights into the seasonal dynamics of dengue in Bangladesh from January 2008 to August 2024, with the highest peak occurring in 2023 at nearly 80,000 cases. The seasonal pattern is evident, with most cases recorded between July and November, and autocorrelation analyses confirming that dengue outbreaks are cyclical, peaking every six months. The SARIMA model (SARIMA (2,1,2) (1,1,1) [6]) effectively captures these seasonal trends, although some autocorrelation remains in the residuals, suggesting room for further refinement. The model predicts recurring peaks in dengue cases from 2024--2027, with significant periodic oscillations. Weather variables, temperature, humidity, and wind speed, were found to influence dengue outbreaks, with increasing case counts. These findings highlight the complex interactions between climate and dengue transmission. Although the Poisson regression models reinforce these relationships, the model’s accuracy could be enhanced by addressing the observed autocorrelation and integrating additional factors such as urbanization. Future studies should focus on refining predictive models, exploring the impacts of climate change, and incorporating socioeconomic variables to develop comprehensive dengue control strategies. Additionally, integrating real-time weather data into early warning systems may improve outbreak preparedness and public health responses.

## Data Availability

All data produced in the present study are available upon reasonable request to the authors

## Abbreviations

DF: Dengue Fever
SARIMA: Seasonal Autoregressive Integrated Moving Average
RMSE: Root Mean Squared Error
IRR: Incidence Rate Ratio
CI: Confidence Interval
ADE: Antibody-dependent enhancement
ARIMA: Autoregressive Integrated Moving Average
SVM: Support vector machines
ANN: Artificial neural networks
HMM: Hidden Markov models
IEDCR: Institute of Epidemiology, Disease Control and Research
BMD: Bangladesh Meteorological Department
AR: Autoregression
MA: Moving Average
MPR: Multivariate Poisson regression
VIF: Variance inflation factors
ACF: Autocorrelation function
PACF: Partial Autocorrelation function
MAE: Mean Absolute Error
SD: Standard deviation
MAPE: Mean absolute percentage error

## Authors Contribution

K.E.A. and M.J.A.: Conceptualization, investigation, data curation, formal analysis and interpretation of data, writing—original draft, writing—review and editing, and project administration, R.C., M.I.H.B., and P.B.: Data collection, writing—original draft, M.A.R., and T.T.M.: Data curation, and D.H.: Data validation and writing— review, editing, and writing. All the authors read the full manuscript and agreed for publication.

## Ethical approval and consent to participate

Not Applicable

## Consent for publication

Not Applicable

## Availability of data and materials

The datasets used and/or analyzed during the current study are available from the corresponding author upon reasonable request.

## Supplementary materials

Supplementary table 1 Meteorological data and dengue cases according to date (Excel data)

## Competing of interests

The authors declare that they have no competing interests.

## Funding

There was no funding for this study.

## Acknowledgment

The authors also thank all the members of the Association of Coding, Technology, and Genomics (ACTG), Sher-e-Bangla Agricultural University (SAU), Dhaka, Bangladesh, who helped to conduct the research. We also appreciate the Directorate General of Public Health and Services, Government of Bangladesh, for providing the required data and access to other necessary information.

## Notes

### Competing Interest Statement

The authors have declared no competing interest.

### Funding Statement

This study did not receive any funding

